# Dissecting the genetic overlap of education, socioeconomic status, and mental health

**DOI:** 10.1101/2020.01.09.20017079

**Authors:** F. R. Wendt, G. A. Pathak, T. Lencz, J. H. Krystal, J. Gelernter, R. Polimanti

**Affiliations:** Department of Psychiatry, Yale School of Medicine, New Haven, CT 06511, USA; VA CT Healthcare Center, West Haven, CT 06516, USA; Division of Psychiatry Research, The Zucker Hillside Hospital, Glen Oaks, NY 11004, USA; Department of Psychiatry, Zucker School of Medicine at Hofstra/Northwell, Hempstead, NY 11549, USA; Institute for Behavioral Science, Feinstein Institute for Medical Research, Manhasset, NY 11030, USA; Departments of Genetics and Neuroscience, Yale University School of Medicine, New Haven, CT 06510, USA

## Abstract

Socioeconomic status (SES) and education (EDU) are phenotypically associated with psychiatric disorders and behavior. It remains unclear how these associations influence the genetic risk for mental health traits and EDU/SES individually. Using information from >1 million individuals, we conditioned the genetic risk for psychiatric disorders, personality traits, brain imaging phenotypes, and externalizing behaviors with genome-wide data for EDU/SES. Accounting for EDU/SES significantly affected the observed heritability of psychiatric traits ranging from 2.44% h^2^ decrease for bipolar disorder to 29.0% h^2^ decrease for Tourette syndrome. Neuroticism h^2^ significantly increased by 20.23% after conditioning with SES. After EDU/SES conditioning, novel neuronal cell-types were identified for risky behavior (excitatory), major depression (inhibitory), schizophrenia (excitatory and GABAergic), and bipolar disorder (excitatory). Conditioning with EDU/SES also revealed unidirectional causality between brain morphology and mental health phenotypes. Our results indicate genetic discoveries of mental health outcomes may be limited by genetic overlap with EDU/SES.

## Introduction

Education (EDU) and socioeconomic status (SES) are risk or protective factors for traits related to mental health and disease (1, 2). Social position has been repeatedly correlated with mood, anxiety, and substance use related disorders, while EDU phenotypes such as *educational attainment, math ability*, and *fluid intelligence* are overall protective factors for development of neurological and psychiatric conditions (2). They are epidemiologically correlated, but the specific EDU and/or SES phenotypes used in epidemiological studies clearly account in part for observed differences between groups (3). It is therefore imperative to understand how EDU and SES phenotypes influence what we understand about human health and disease.

Genome-wide association studies (GWAS) are powerful hypothesis-free genetic studies for detecting risk loci (e.g., single nucleotide polymorphisms (SNPs) or genes) for phenotypes of interest. Their widespread use has led to risk locus discovery underlying thousands of phenotypes across the spectrum of human health and disease, including mental and physical health and disease, personality, anthropometric measures, intelligence, and behavior (4). An observation generated from large-scale GWAS is the widespread presence of pleiotropy; a single SNP (or a set of SNPs) may have a range of relatively small effects on multiple similar or disparate phenotypes. On a genome-wide scale, these pleiotropic effects, detected using GWAS summary data, may be used to determine genetic correlations between phenotypes to putatively identify genetic underpinnings of trait pairs (5).

The EDU phenotypes *educational attainment* and *cognitive performance* have relatively high SNP-heritability: the phenotypic variance explained by genetic information was 40-60% (6) and 21.5% (7), respectively. Socioeconomic status (SES) is defined as the social standing or class of an individual or group, often measured as a combination of education, income, and occupation (8). SES phenotypes such as *household income* and *Townsend deprivation index* (i.e., measure of SES based on whether individuals own their homes, their employment status, their access to a vehicle, and whether or not individuals share living accommodations with others) are significantly heritable and show strong genetic correlation with EDU traits (9). Additionally there is pleiotropy of genetic risks between EDU/SES and a range of mental health outcomes (e.g., psychiatric disorders, personality traits, internalizing and externalizing behaviors, social science outcomes, and brain imaging phenotypes) (10, 11).

The epidemiological observations of high genetic correlations between genetic risk for EDU/SES and mental health outcomes (1, 2) raise two critical questions: (1) how might the strong genetic effects of EDU/SES affect our understanding of the overall genetic risk for mental health outcomes? and (2) is there evidence that genetic effects of mental health and disease phenotypes affect our understanding of the overall genetic risk for EDU/SES? The goal of this study was to investigate how the shared genetic effects between the general categories of EDU, SES, and mental health outcome phenotypes influence genetic risk for individual phenotypes within each of these classes.

There are several ways to approach these questions. First, polygenic risk scoring (PRS) (12) is a tempting approach; but PRS using mental health/disease to predict the same or different phenotypes from an independent dataset often explain very little variance in the outcome phenotype (13-15). PRS also cannot detect specific biology underlying each phenotype. Second is multi-trait analysis of GWAS (MTAG), which jointly analyses GWAS summary statistics and adjusts per-SNP effect estimates and association p-values using the strength of the genetic correlation between phenotypes (16). Genetic correlations between EDU/SES and related phenotypes have, however, demonstrable biases from environmental confounders. If genetic correlations involving EDU and SES proxy phenotypes are significantly upwardly biased, an MTAG adjustment of summary statistics may inappropriately correct (i.e., bias) the summary statistics used for this study. To disentangle the complex genetic overlaps between EDU/SES and mental health, we therefore used multi-trait conditioning and joint analysis (mtCOJO), which generates conditioned GWAS summary statistics for each phenotype of interest after correcting for the per-SNP effects of another phenotype (17). The mtCOJO approach is not based on genetic correlation; it is based on the causal relationship between trait pairs inferred by Mendelian randomization (MR). For our phenotypes of interest mtCOJO is an advantageous approach, which, in theory, is independent of the effects of environmental confounders. MR detects causal inferences between trait pairs using non-modifiable risk factors (SNPs) associated with an exposure variable and only associated with an outcome variable through the exposure. Because SNPs are non-modifiable, environmental confounders of the relationship between SNP, exposure, and outcome should not influence MR estimates.

We used the mtCOJO approach to condition mental health outcomes with the per-SNP effects of EDU and SES phenotypes and investigate their underlying biology at multiple levels: (1) risk locus detection, (2) heritability (h^2^), (3) gene-set enrichment, (4) tissue transcriptomic profile enrichment, (5) cell type transcriptomic profile enrichment, (6) phenotype relationships via structural equation modeling and genetic correlation, and (7) latent genetically causal relationships (see flow diagram Fig S1). Our findings identify several cell types and phenotype relationships that were masked by the shared genetic etiology between mental health outcomes and EDU/SES. Furthermore, we demonstrate that the same multi-level analyses of EDU and SES are largely robust to the effects of shared genetic etiology with mental health outcomes.

## Results

### Trait Inclusion

The genetic correlations (r_g_) between EDU (*educational attainment, cognitive performance, highest math class*, and *self-rated math ability*), SES (*household income* and *Townsend deprivation index*), and mental health outcomes (i.e., psychiatric disorders, personality traits, externalizing behaviors, social science outcomes, and brain imaging phenotypes) were detected using the Linkage Disequilibrium Score Regression (LDSC) method (Fig. 1, Table S1, Figs. S2 & S3) (18). Genetic correlations for EDU and SES phenotype categories were analyzed independently to identify brain imaging phenotypes nominally genetically correlated with at least two of the four EDU phenotypes and both SES phenotypes. We detected only two traits genetically correlated with at least two of the four EDU phenotypes: *left insular cortex* (mean r_g_ = −0.122, se = 0.013) and *left subcallosal cortex* (mean r_g_ = −0.106, se = 0.009). These two brain imaging phenotypes were included in EDU conditioning experiments.

**Fig. 1.**
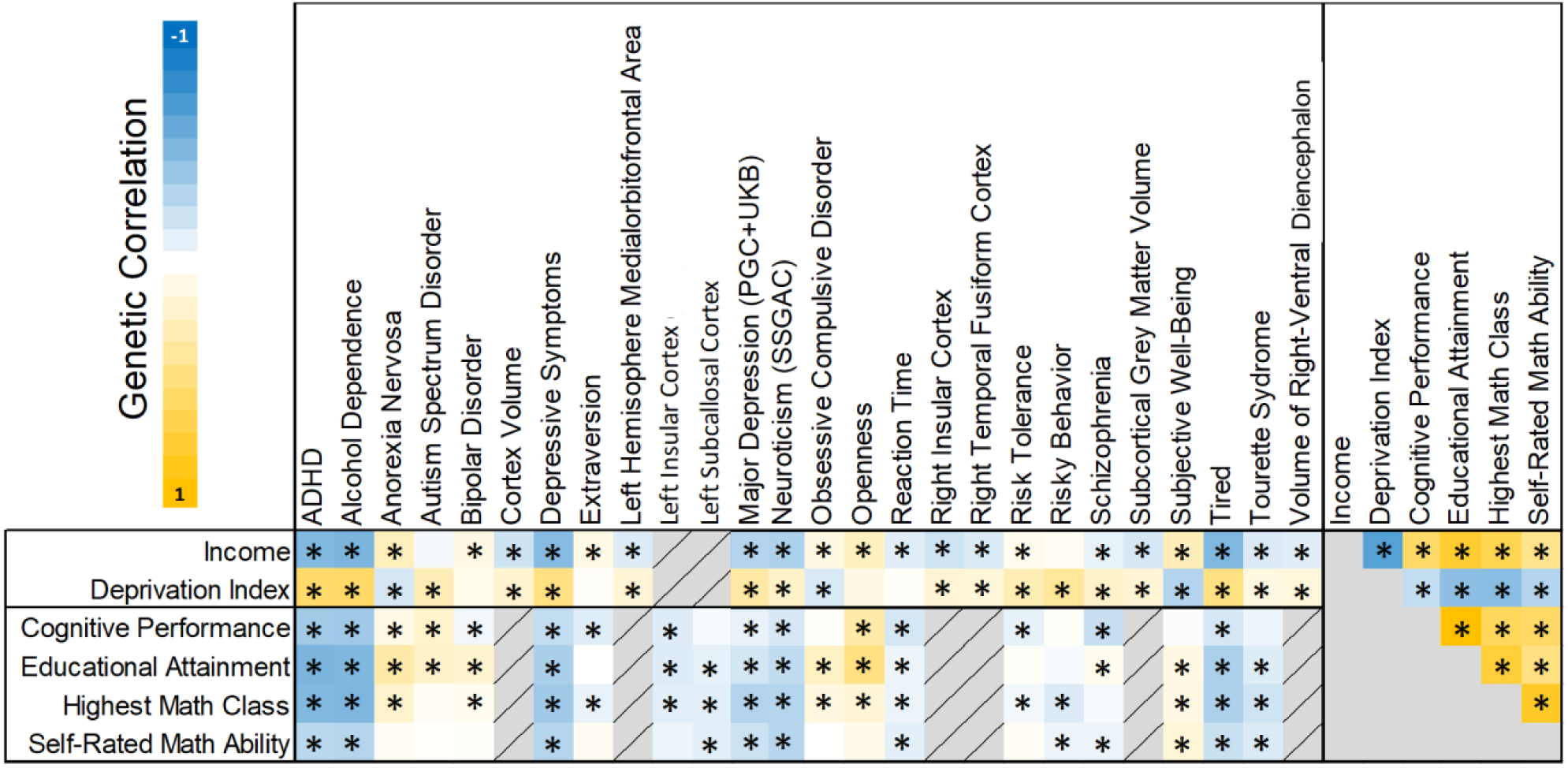
Trait inclusion genetic correlations. Genetic correlation between mental health outcomes, education phenotypes, and socioeconomic status phenotypes. Genetic correlations labeled with an asterisk were at least nominally significant.

Twenty-nine brain imaging phenotypes were genetically correlated with both SES phenotypes. We tested genetic correlation between these 29 brain imaging phenotypes to identify a subset of high heritability traits to include in SES conditioning experiments. We identified six such brain imaging phenotypes (Fig. S2). These are: *cortex volume, left hemisphere medialorbitofrontal area, right insular cortex, right temporal fusiform cortex, subcortical gray matter volume*, and *volume of right-ventral diencephalon*. The SES phenotypes *income* and *deprivation index* are inversely genetically correlated as visible in Fig. 1.

### Conditioning Heritability and Risk Locus Discovery

We tested the effects of conditioning on observed-scale heritability (h^2^) using LDSC (18). Psychiatric disorders were most sensitive to shared genetic etiology with EDU/SES phenotypes. Except for *major depressive disorder* (*MDD*), *anxiety*, and *posttraumatic stress disorder* (*PTSD*), conditioning reduced the h^2^ for all psychiatric disorders relative to their original estimates (h^2^ decrease ranged from 2.44% ± 0.187 for *bipolar disorder* (original h^2^ = 4.39%; highest conditioned h^2^ = 2.22%, se = 0.460, p = 5.67×10^−65^; lowest conditioned h^2^ = 1.70%, se = 0.440, p = 4.05×10^−80^) to 29.0% ± 0.105 for *Tourette syndrome* (original h^2^ = 35.6%; highest conditioned h^2^ = 6.72%, se = 0.770, p = 2.61×10^−18^; lowest conditioned h^2^ = 6.43%, se = 0.730, p = 1.27×10^−18^); Fig. 2A). *Tourette syndrome* exhibited the largest decrease in h^2^ after conditioning with the effects of EDU/SES phenotypes (*Tourette syndrome* mean p_diff_ compared to original h^2^ = 2.24×10^−11^, se = 4.42×10^−12^). Conversely, two phenotypes exhibited significant increases in h^2^ after conditioning with EDU/SES phenotypes: *neuroticism* (highest conditioned h^2^ = 20.2%, se = 0.630, p = 3.08×10^−226^; lowest conditioned h^2^ = 18.1%, se = 0.590, p =2.35×10^−207^) and *subjective well-being* (highest conditioned h^2^ = 3.65%, se = 0.220, p = 8.11×10^−62^; lowest conditioned h^2^ = 3.34%, se = 0.220, p =4.67×10^−52^).

**Fig. 2.**
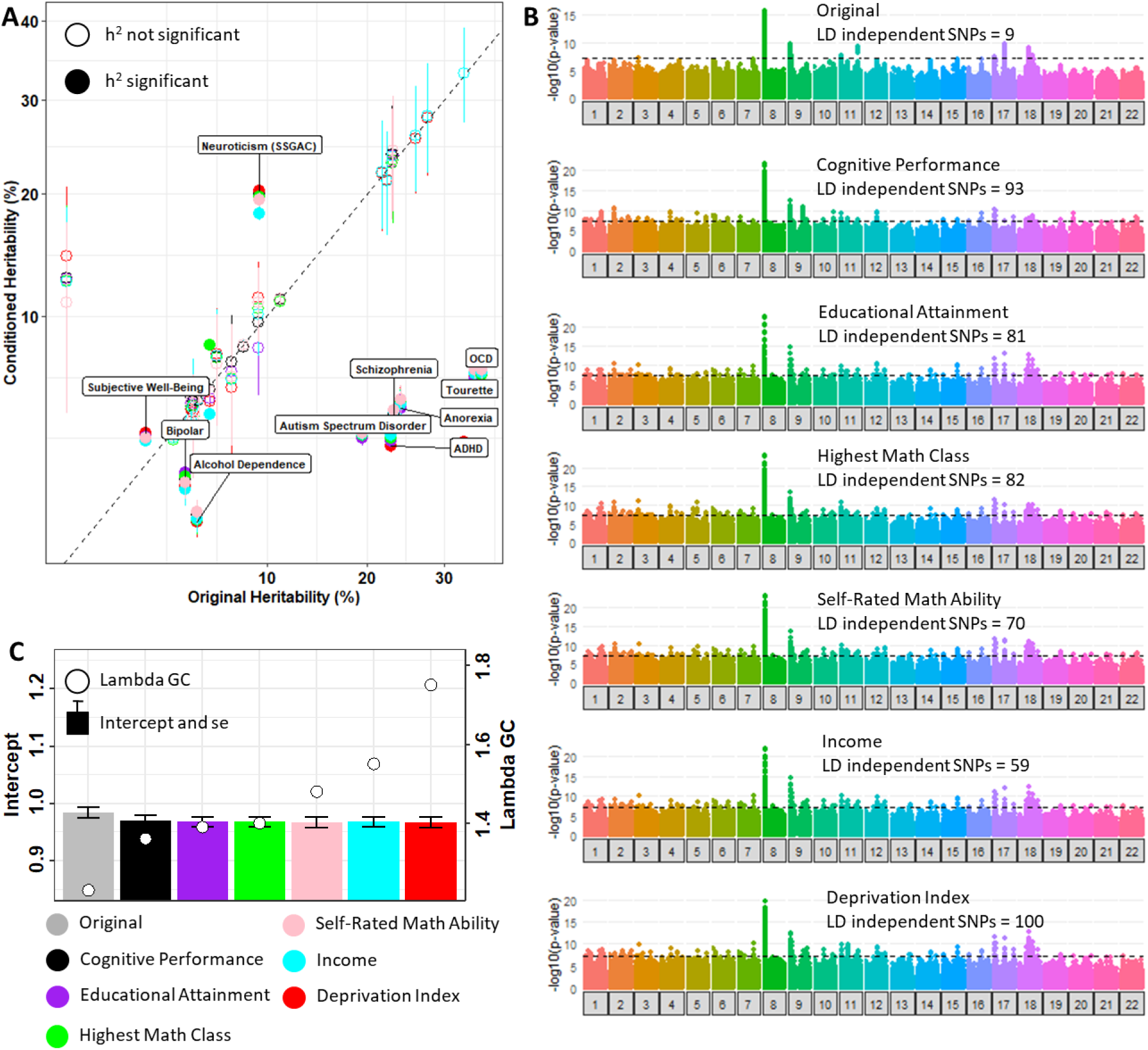
Heritability (h_2_) changes and risk locus discovery. **(A)** Observed-scale h^2^ changes of mental health outcomes after conditioning with education and socioeconomic status phenotypes. **(B)** Manhattan plots for neuroticism (SSGAC) before and after conditioning with education and socioeconomic status phenotypes. **(C)** Evidence that neuroticism (SSGAC) locus discovery is due to increased detection of polygenicity rather than exacerbated effects of population substructure.

Conditioning the *neuroticism* GWAS (original h^2^ = 9.41%) with EDU/SES phenotypes revealed several novel, confirmed known LD-independent risk loci, and increased heritability (range = 59 loci (*neuroticism* conditioned with *income*) to 100 loci (*neuroticism* conditioned with *deprivation index*; Fig. 2B). We observed an increase in the association signal in the *neuroticism* GWAS with the strongest effects observed after conditioning with SES phenotypes *income* (lambda GC = 1.36; intercept = 0.971, se = 0.009) and *deprivation index* (lambda GC = 1.75; intercept = 0.967, se = 0.009; Fig. 2C). This increase was not related to an increase in the potential bias of population stratification (there was no significant change in the LDSC intercept, p > 0.05), supporting that the observation was attributable to the increased detection of valid *neuroticism* polygenic signals. Using a physical proximity single-SNP-single-gene based annotation of conditioned *neuroticism* genomic risk loci, the top gene sets included Gene Ontology (GO) biological process synaptic signaling (enrichment FDR = 1.5×10^−4^), GO cellular component synapse part (enrichment FDR = 5.40×10^−4^), and Kyoto Encyclopedia of Genes and Genomes (KEGG) dopaminergic synapses (enrichment FDR = 0.046).

The significant increase in h^2^ for GWAS of *subjective well-being* (original h^2^ = 2.50%) uncovered a 5.7 kb genomic risk locus on chromosome 7 (minimum genome-wide significant p-value = 1.45×10^−8^) which maps to the α_2_δ_1_ subunit of calcium voltage-gated channel (*CACNA2D1*). The protein encoded by *CACNA2D1* has been implicated in familial epilepsy and intellectual disability pedigrees but to our knowledge has not been implicated in genome-wide studies of these phenotypes (19, 20).

### Tissue-Type Transcriptomic Profile Enrichment Differences

After conditioning with GWAS of EDU/SES phenotypes, *schizophrenia* was the only mental health outcome demonstrating significant changes in tissue transcriptomic profile enrichment. Compared to original *schizophrenia* brain tissue transcriptomic profile enrichments, all conditioned *schizophrenia* brain tissue GTEx annotations, with the exception of c1 cervical spinal cord, had significantly decreased enrichments (Fig. S4). The maximum decrease was observed after conditioning *schizophrenia* with the EDU phenotype *educational attainment* (average beta decrease for all brain tissue annotations = 0.038 ± 0.004). After conditioning with EDU and SES phenotypes, the cerebellum and cerebellar hemisphere GTEx annotations remained the most enriched in the *schizophrenia* GWAS (original cerebellum enrichment = 0.080, p = 1.76×10^−22^; original cerebellar hemisphere enrichment = 0.077, p = 1.28×10^−22^; mean conditioned cerebellum enrichment = 0.047 ± 0.001, FDR < 0.05; mean conditioned cerebellar hemisphere enrichment = 0.047 ± 0.001, FDR < 0.05). After adjusting for the effects of *cognitive performance* and *educational attainment* and correcting for multiple testing, we uncovered enrichment of skeletal muscle tissue transcriptomic profiles in the *schizophrenia* GWAS (original skeletal muscle enrichment = 0.009, p = 0.135; skeletal muscle enrichment conditioned with *educational attainment* = 0.010, p = 0.032; skeletal muscle enrichment conditioned with *cognitive performance* = 0.011, p = 0.024) (21).

### Cell-Type Transcriptomic Profile Discoveries

Cell-type transcriptomic profile enrichments were evaluated in two ways: (1) assess differences in within-data-set cell-type enrichments before and after conditioning with EDU/SES (based on MAGMA cell-type enrichment Step 1 (22)) and (2) assess the effects of conditioning on the detection of conditionally independent proportionally significant (PS) cell type enrichments (based on MAGMA cell-type enrichment Step 3 (22)). PS cell-types are those whose genetic signals could be differentiated from one another. PS values ≥ 0.80 indicate independent genetic signals relative to a second cell type. We then used genes whose expression profiles define the excitatory (Ex) and inhibitory (In) cell types of PsychENCODE (23) to perform gene set enrichment analyses of GO and KEGG gene sets.

There were no differences in cell-type transcriptomic profile enrichments for mental health outcomes (MAGMA cell-type Step 1) after conditioning with EDU/SES; however, we discovered several PS cell-type pairs not detected in the unconditioned GWAS for (1) *risky behavior*, (2) *MDD*, and (3) *schizophrenia* (MAGMA cell-type Step 3). These PS cell-type findings and relevant gene set results for are described in detail below.

In unconditioned GWAS of *risky behavior*, there were no PS cell-type enrichments After conditioning with the EDU phenotypes *cognitive performance* and *educational attainment*, human cortex fetal quiescent and Ex2 were conditionally independent from one another (*risky behavior* conditioned with *cognitive performance* Ex2 β = 0.035, p = 7.48×10^−4^, PS = 1.37; fetal quiescent β = 0.023, p = 0.032, PS = 1.82; *risky behavior* conditioned with *educational attainment* Ex2 β = 0.034, p = 0.001, PS = 1.38; fetal quiescent β = 0.024, p = 0.030, PS = 1.77; Fig 3A). Ex2 neurons also were detected in the *risk tolerance* GWAS after conditioning with *educational attainment*, but this signal could not be distinguished from hippocampal CA1 subfield cells. The genes that define the Ex2 cell type were enriched in nervous system development (GO:0007399; enrichment FDR = 3.70×10^−4^) and eye development (GO:001654; enrichment FDR = 6.30×10^−4^) gene sets.

**Fig. 3.**
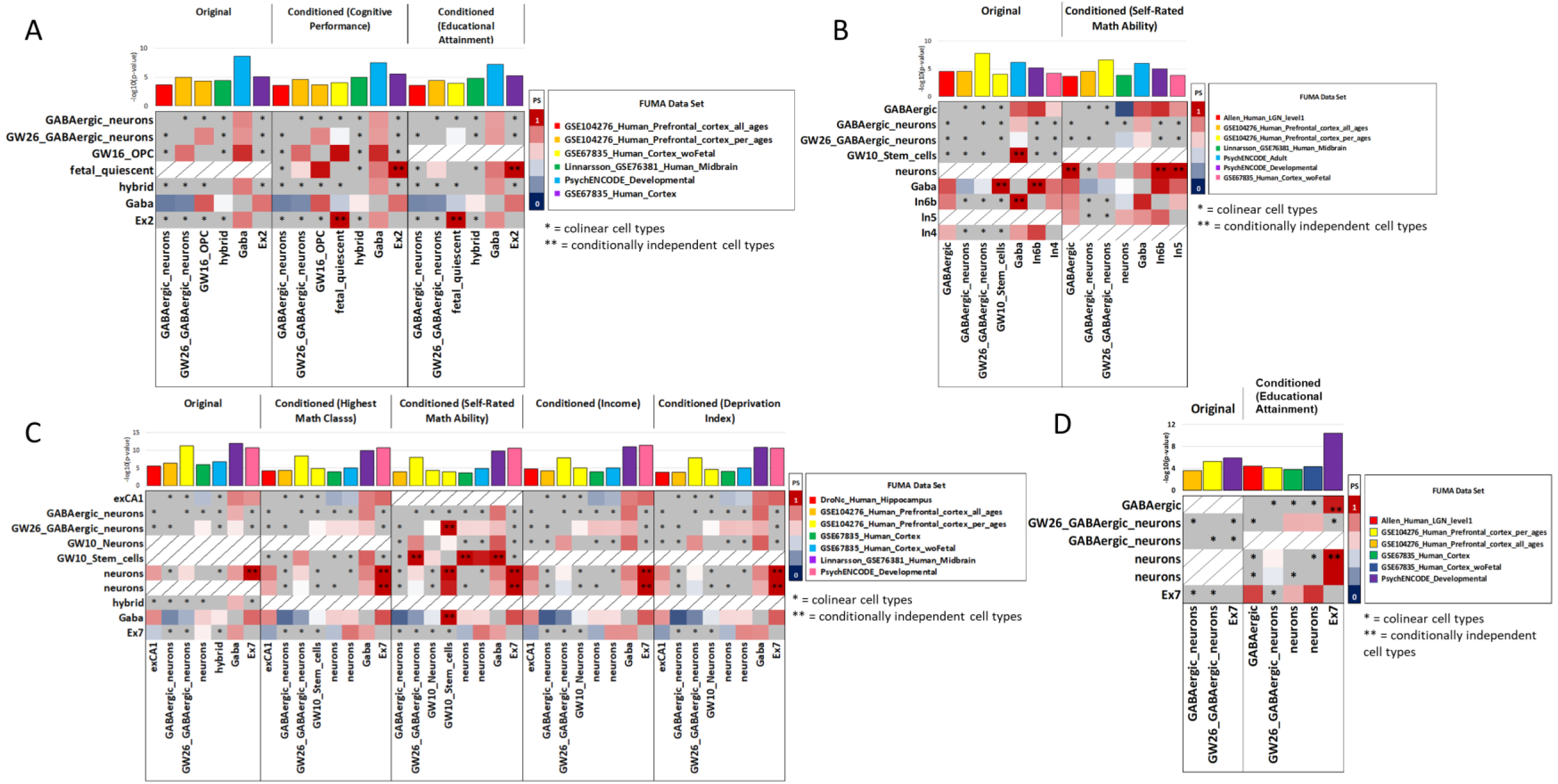
Cell-type transcriptomic profile enrichments underlying mental health outcomes. Cross-data-set proportionally significant (PS) and conditionally independent (i.e., genetic signatures of cell-type pairs are distinguishable) cell-type transcriptomic profile enrichments underlying unconditioned and conditioned GWAS for **(A)** risky behavior, **(B)** major depression, **(C)** schizophrenia, and **(D)** bipolar disorder. The human cell-type data sets from FUMA are labeled individually for each panel using different colors; cell types in the x and y directions are conditionally independent signals from within-data-set analysis performed in FUMA (cell-type enrichment step 2 (22)). Genetic signals from colinear cell types labeled with a single asterisk could not be differentiated from one another in FUMA.

The unconditioned *MDD* GWAS exhibited cell-type transcriptomic profile enrichments between adult GABAergic neurons, In6b, and gestational week 10 (GW10) stem cells. After conditioning with *self-rated math ability*, the genetic signal from human midbrain neurons was conditionally independent from lateral geniculate nucleus (LGN) GABAergic neurons (β relative to midbrain neurons = 0.041, p = 0.002, PS = 0.822; Fig 3B), In6b neurons (β relative to midbrain neurons = 0.517, p = 6.59×10^−6^, PS = 0.969), and In5 neurons (β relative to midbrain neurons = 0.039, p = 5.26×10^−5^, PS = 0.813). The gene expression profiles of these cell types implicate the neurotransmitter transport (GO:0007269; enrichment FDR = 0.003) and locomotory behavior (GO:0007626; enrichment FDR = 0.015) gene sets in *MDD* psychopathology.

The cell-type transcriptomic profiles underlying *schizophrenia* initially highlighted the role of Ex7 and human cortical neurons with conditionally independent genetic signals. After conditioning the *schizophrenia* GWAS with *self-rated math ability*, we uncovered conditionally independent PS genetic signals from GW26 GABAergic neurons and GW10 stem cells (GABAergic neuron β = 0.046, p = 9.46×10^−9^, PS = 1.00; GW10 stem cell β = 0.031, p = 1.04×10^−4^, PS = 1.00; Fig 3C and 3D). Importantly, the independent genetic signals of Ex7 and human cortical neurons persisted after conditioning the *schizophrenia* GWAS with EDU and SES phenotypes. There were no conditionally independent PS cell-type signals in the unconditioned GWAS, but the GWAS of *bipolar disorder* conditioned with *educational attainment* revealed PS genetic signals from (1) Ex7 (β relative to GABAergic neurons from the lateral geniculate nucleus (LGN) = 0.043, p = 1.50×10^−10^, PS = 0.952 and Ex7 beta relative to LGN human cortical neurons = 0.035, p = 5.46×10^−6^, PS = 0.999), (2) LGN GABAergic neurons (β relative to Ex7 = 0.045, p = 1.42×10^−4^, PS = 0.871), and (3) human cortical neurons (β relative to Ex7 = 0.001, p = 0.044, PS = 0.904) but could not distinguish genetic signals between the LGN GABAergic and human cortical neuron cell types. The genes contributing to the Ex7 cell type were enriched in gene sets related to nervous system processes (GO:0050877; enrichment FDR = 0.014) and synaptic signaling (GO:0099536; enrichment FDR = 0.023).

### Correlative, Latent, and Causal Relationships between Mental Health Outcomes

Genetic correlations were assessed between all mental health outcomes after conditioning with each EDU and SES phenotype. Though small changes in genetic correlation magnitude were observed, the mental health outcome genetic correlations largely persisted even after conditioning with EDU/SES (Fig. S5). Two mental health outcomes, however, demonstrated significant changes in their genetic correlations after conditioning: (1) genetic correlations with *neuroticism* and (2) genetic correlations with *volume of the right-ventral diencephalon*. The genetic correlations between conditioned *neuroticism* and (1) *MDD* (original r_g_ = 0.732, mean conditioned r_g_ = 0.574 ± 0.008), (2) *subjective well-being* (original r_g_ = −0.718, mean conditioned r_g_ = −0.522 ± 0.009), and (3) *tiredness* (unconditioned r_g_ = 0.638, mean conditioned r_g_ = 0.490 ± 0.017) were at least nominally significant, and were in each case significantly lower than the unconditioned relationship. Unconditioned *right-ventral diencephalon volume* was significantly genetically correlated with *subcortical gray matter volume* (unconditioned r_g_ = 0.620, p = 8.77×10^−15^) and *schizophrenia* (unconditioned r_g_ = 0.134, p = 0.009). After conditioning with *income*, the genetic correlation between *volume of the right-ventral diencephalon* and (1) *subcortical gray matter volume* persisted (conditioned r_g_ = 0.612, p = 1.44×10^−14^), (2) *schizophrenia* switched directions and remained significant (conditioned r_g_ = −0.120, p = 0.011, p_diff_ = 2.72×10^−4^), and (3) *risk tolerance* became significant (conditioned r_g_ = −0.123, p = 0.044). Conversely, after conditioning with the effects of *deprivation index*, the genetic correlation between *volume of the right-ventral diencephalon* and (1) *subcortical gray matter volume* was no longer significant (conditioned r_g_ = −0.076, p = 0.315), (2) *schizophrenia* increased in magnitude (conditioned r_g_ = 0.198, p = 5.20×10^−12^, p_diff_ = 0.294), and (3) several additional phenotypes become at least nominally significant (conditioned r_g_ with *autism spectrum disorder* (*ASD*) = 0.478, p = 3.40×10^−30^; with *bipolar disorder* = 0.181, p = 2.93×10^−8^; with *risky behavior* = 0.418, p = 1.50×10^−59^; with *subjective well-being* = −0.363, p = 1.83×10^−17^; and with *tiredness* = 0.343, p = 1.82×10^−20^; Fig. S5).

Genomic Structural Equation Modeling (GenomicSEM) was used to identify how unconditioned and conditioned mental health outcomes relate to a latent unobserved genetic factor connecting them (Fig. 4). In unconditioned models, exploratory factor analysis (EFA) identified a two-factor model as best suited to explain the relationships among mental health outcomes. In confirmatory factor analysis (CFA), these two latent factors generally highlight relationships between all psychiatric disorders and brain imaging phenotypes (F1) and *anxiety, MDD, depressive symptoms*, and *neuroticism* (F2). The correlation between unconditioned F1 and F2 was 0.14. After conditioning with *highest math class, self-rated math ability*, and *deprivation index*, the GWAS of *neuroticism* and *MDD* were no longer major contributors to the same factor. Conditioned F1 had major contributions from *MDD* (mean loading = 0.611 ± 0.005) and *depressive symptoms* (loading = 0.538 ± 0.098) while conditioned F2 had major contributions from *neuroticism* (loading = 0.877 ± 0.080) and *anxiety* (loading = 0.658 ± 0.009). Interestingly, after conditioning with the SES phenotype *income*, the SEM best-fit converged on a single common factor between all mental health outcomes with major contributions from *MDD* (loading = 0.808, se = 0.068) and *depressive symptoms* (loading = 0.831, se = 0.022).

**Fig. 4.**
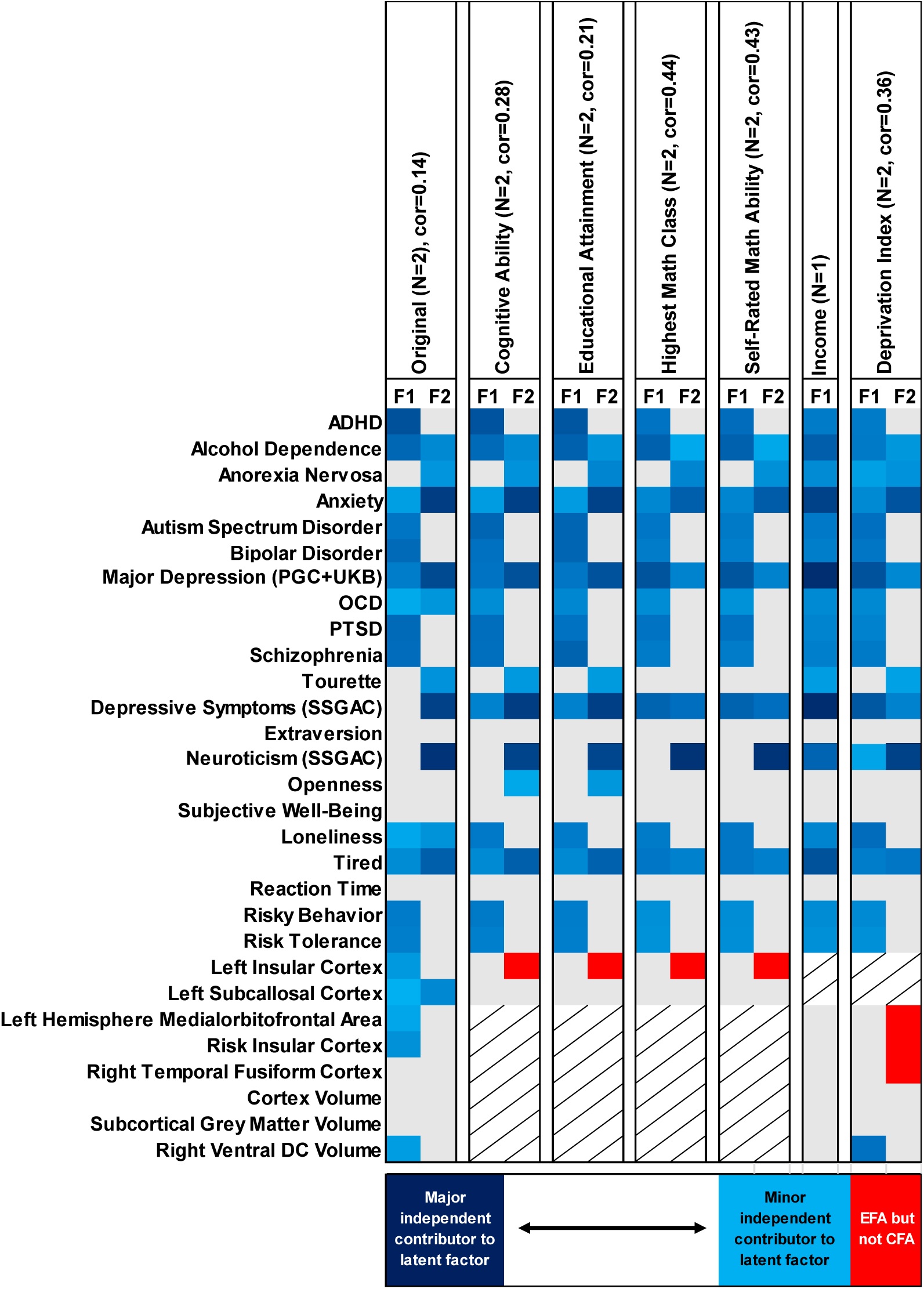
Trait loading onto latent factors. Genomic structural equation modeling of mental health outcomes before and after conditioning with education and socioeconomic status phenotypes. Each column shows the confirmatory factor analysis (CFA) loading value (blue shading indicating that a trait is a major contributor to the latent factor and blue tinting indicating that a trait is a minor independent contributor to the latent factor) for each mental health outcome (in the y direction) into one of two factors (F1 and F2) from exploratory factor analysis (EFA). Grey boxes indicate that a given trait was not predicted to load onto a given factor column. Red boxes indicate that the trait was predicted by EFA to load onto a factor but did not independently load during CFA.

Latent Causal Variable (LCV) analyses were used to detect causal relationships between trait pairs that are independent of the genetic correlations between them (24). Considering only the unconditioned mental health outcomes, one trait pair exhibited significant genetic causality proportion (gĉp): *left subcallosal cortex*→*obsessive compulsive disorder* gĉp = 0.167, p = 4.54×10^−6^ (Table 1 and Fig. 5). This partial causal relationship did not survive conditioning; however, thirteen unique trait pairs demonstrated significant gĉp after conditioning both traits with an EDU or SES phenotype (Table 1). Most notable were those causal relationships involving brain imaging phenotypes which became significant after conditioning with EDU phenotypes: (1) *extraversion*→*left subcallosal cortex* (mean gĉp = 0.188 ± 0.107, 1.23×10^−13^<p-values<1.83×10^−6^) after conditioning with *educational attainment, highest math class*, and *self-rated math ability*, (2) *left subcallosal cortex*→*subjective well-being* (mean gĉp = 0.745 ± 0.009, 1.45×10^−9^<p-values<1.16×10^−8^) after conditioning with *cognitive performance, educational attainment*, and *highest math class*, (3) *openness*→*left insular cortex* (mean gĉp = 0.296 ± 0.050, 2.54×10^−23^<p-values<3.63×10^−8^) after conditioning with *cognitive performance, highest math class*, and *self-rated math ability*. These average gĉp estimates represent only Bonferroni significant relationships; however, each trait pair listed was nominally significant after conditioning with all other EDU and SES phenotypes but not significant in the unconditioned experiment (Table 1).

**Fig 5.**
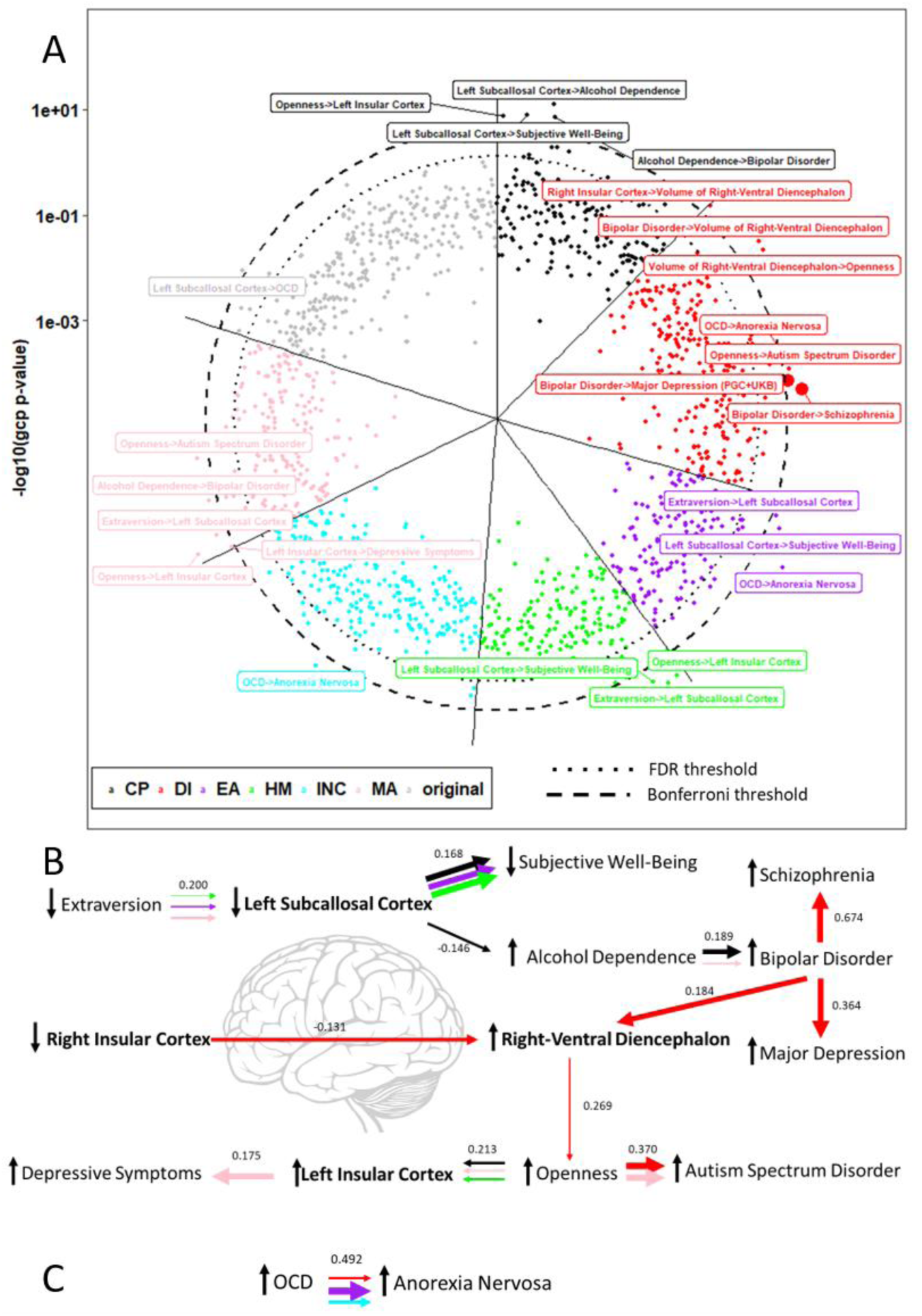
Causal relationships masked by education and socioeconomic status effects. Latent Causal Variable (LCV) results detecting causal relationships between the genetic risk for two mental health outcomes. **(A)** Colors are used to indicate original trait pair and conditioned trait pair LCV results. LCV genetic causality proportions surviving multiple testing correction are labeled. Large data points indicate significantly different LCV genetic causality proportions relative to the unconditioned estimate. **(B)** Summary of the causal relationship (derived from **A**) network originating from brain imaging phenotypes (bolded text). **(C)** Causal relationships with no evidence of brain imaging phenotype connection in the current study (derived from **A**). In **B** and **C**, horizontal arrow thickness indicates the size of the estimated causal relationship between the two traits on either end of the arrow while the direction of each arrow indicates the direction of causal effect; the color of each arrow indicates the education or socioeconomic status phenotype used to condition each trait of a trait pair; mean genetic correlations from LCV are included above each set of horizontal arrows.

The EDU/SES phenotype which revealed the most latent causal relationships between mental health outcomes was *Townsend deprivation index*. Conditioning with this phenotype revealed 7/13 causal relationships, most of which involved *bipolar disorder* or the *volume of the right-ventral diencephalon*.

### Effects of Mental Health Outcomes on Education and Socioeconomic Status

We evaluated how shared etiology between mental health outcomes and EDU/SES phenotypes might influence our understanding of the genetics of EDU/SES. After conditioning each EDU and SES phenotype with individual mental health outcomes, all six EDU/SES phenotypes were significantly heritable and maintained high genetic correlation with their unconditioned equivalents (Fig. S6). *Attention deficit hyperactivity disorder* (*ADHD*) and *tiredness* had the most substantial influence on the h^2^ of EDU and SES phenotypes, both of which significantly reduced the h^2^ of EDU/SES phenotypes relative to their original h^2^ estimates. Next, each EDU and SES phenotype was simultaneously conditioned with all genetically correlated mental health outcomes to evaluate how robust EDU and SES phenotypes are to genetic etiology shared with many phenotypes. After conditioning with all mental health outcomes, the EDU/SES phenotypes *educational attainment, income*, and *self-rated math ability* maintain significant h^2^. Conditioned *educational attainment* (mean r_g_ = 0.999 ± 0.006), *income* (mean r_g_ = 0.988 ± 0.025), and *self-rated math ability* (mean r_g_ = 0.987 ± 0.033) were highly genetically correlated with their unconditioned phenotypes.

Gene set, tissue transcriptomic profile, and cell-type transcriptomic profile enrichments were generally robust to the effects of individual mental health outcomes (Fig S7). After conditioning with *MDD*, the GWAS of *self-rated math ability* demonstrated a significant decrease in brain cortex tissue transcriptomic profile enrichment (conditioned frontal cortex (BA9) enrichment = 0.106, p = 1.61×10^−23^; conditioned cortex enrichment = 0.108, p = 4.48×10^−23^; conditioned anterior cingulate cortex (BA24) enrichment = 0.106, p = 6.12×10^−21^). We then evaluated these enrichment changes in the EDU and SES phenotypes which maintained significant h^2^ after conditioning with all mental health outcomes (*educational attainment, income*, and *self-rated math ability*). Conditioned *educational attainment* demonstrated changes in enrichment of neurogenesis (p_diff_ = 1.43×10^−8^) and neuron differentiation gene sets (p_diff_ = 1.61×10^−7^) such that they were no longer associated with the GWAS of *educational attainment*. Conditioned EDU phenotypes *educational attainment* (9.57×10^−17^<p_diff_<2.14×10^−8^) and *self-rated math ability* (4.35×10^−14^<p_diff_<4.21×10^−12^) both demonstrated decreased enrichment of cerebellar and cortical tissue transcriptomic profiles. Cell-type transcriptomic profile enrichment changes were generally unique to each phenotype: *educational attainment* demonstrated decreased enrichment of inhibitory (In4, p_diff_ = 6.38×10^−14^) and GABAergic (p_diff_ = 8.03×10^−11^) neurons, *self-rated math ability* demonstrated decreased enrichment of excitatory (Ex1 (p_diff_ = 7.37×10^−11^), Ex2 (p_diff_ = 1.27×10^−10^), Ex4 (p_diff_ = 9.96×10^−10^), Ex5 (p_diff_ = 7.27×10^−10^), and Ex7 (p_diff_ = 1.77×10^−10^)) neurons, and *income* demonstrated changes in excitatory (Ex1 (p_diff_ = 0.004), Ex4 (p_diff_ = 0.005), Ex5 (p_diff_ = 0.001), and Ex9 (p_diff_ = 0.002)), inhibitory (In4; p_diff_ 0.001), and GW26 GABAergic (p_diff_ = 4.67×10^−4^) neurons. Consistent with our discoveries regarding cell types underlying mental health outcomes, cell type enrichment changes underlying conditioned EDU and SES phenotypes reflect changes in nervous system development, synaptic signaling, and eye development gene ontologies.

In a structural equation model assuming a single factor connecting all EDU and SES phenotypes, *income* was the only trait that did not load independently of all other EDU/SES proxies (loading = −0.781; Fig. S8). EFA of EDU and SES phenotypes revealed a two-factor model with major contributions from *educational attainment* (F1 loading = 0.990, se = 0.015) and *self-rated math ability* (F2 loading = 0.996, se = 0.046). The correlation between F1 and F2 was 0.426 and was mediated by *cognitive performance* (F1 loading = 0.429, se = 0.018; F2 loading = 0.292, se = 0.027) and *highest math class* (F1 loading = 0.603, se = 0.024; F2 loading = 0.466, se = 0.029). Notably, the SES phenotype *income* also did not load onto either factor independently of the other EDU and SES phenotypes in EFA.

After conditioning with all mental health outcomes, we reanalyzed the structural equation models using only those EDU/SES phenotypes with significant h^2^ (*educational attainment, income*, and *self-rated math ability*). Assuming a single factor (as well as EFA and CFA), *educational attainment* remained a major contributor to the model (common factor loading = 0.986, se = 0.127). In CFA of conditioned phenotypes, the loading value for *income* significantly increased relative to its original loading value (conditioned common factor loading = 0.118, se = 0.253, p_diff_ = 0.018) such that *income* independently loaded onto the same common factor as *educational attainment* and *self-rated math ability*.

## Discussion

EDU and SES are important influences on numerous mental health and disease phenotypes, but it has been difficult to determine the extent to which this is so, and the biological nature of the relationship. How much of the genetic risk for *schizophrenia*, for example, is caused by reduced *educational attainment*? Or how much of the risk for schizophrenia reflects a shared biology with the predisposition to *educational attainment*? These are important questions to answer if we are to understand the biology of both kinds of traits. To get at this question, we conditioned one on the other, and thereby statistically removed its effects, and then asked the question, “how much of the heritable risk for that trait remains?” In most cases, SES/EDU accounted for some of the genetic variance in the mental health or disease phenotype and adjusting for SES/EDU reduced the strength of the association with the heritable risk for that disorder. However, in a few cases (depression, anxiety, neuroticism, PTSD, and subjective well-being) adjusting for SES/EDU either increased or did not change these associations. In the space below, we present a framework for interpreting these complexly of these findings.

### Mental Health Outcomes

The biology underlying psychiatric disorders was most affected by shared genetic etiology with EDU/SES proxies, as evidenced by significant decreases in h^2^ for all psychiatric disorder except *MDD, anxiety*, and *PTSD* when conditioned on these traits. Conversely, conditioning the *neuroticism* and *subjective well-being* GWAS revealed additional risk loci that were not detected in the original GWAS. Using an independent method, Turley, et al. observed similar information gain (16); however, we demonstrated that this information gain is due to polygenicity rather than population substructure. That is, we believe this demonstrated novel biology, as opposed to the underlying population genetics phenomena. Unlike Turley, et al., we do not report comparable risk locus gain with *subjective well-being*.

In structural equation models of mental health outcomes, *neuroticism* and *MDD* originally loaded onto the same common factor. After conditioning with *highest math class, self-rated math ability*, and *deprivation index*, the loadings of *neuroticism* and *MDD* separate, suggesting that their unique biology may be distinguishable. This is consistent with our observation of ameliorated genetic correlation between these two phenotypes due to conditioning. Hill, et al., described two factors of *neuroticism*, one of which aligns more closely with anxiety and tension phenotypes and the second of which aligns more closely with worry and vulnerability phenotypes (25). With GenomicSEM, we support these claims: *neuroticism* loads onto the same common factor as *anxiety* while *MDD* aligns with *depressive symptoms* and *loneliness*. We demonstrate here that *neuroticism* and *MDD* are highly positively genetically correlated in their unconditioned versions. Based on the present results, we hypothesize that conditioning these phenotypes with EDU and SES reveals unique genetic architectures of these phenotypes. We demonstrate that after conditioning with EDU/SES, general neuroticism appears more similar to the Hill, et al. Anxiety/Tension phenotype. Lastly, our GenomicSEM data mirror those genetic correlation results from Hill, et al., adding weight to our observed two-factor model (25).

Cell type transcriptomic profile enrichments underlying the GWAS of mental health outcomes were robust to the effects of EDU and SES phenotypes, but we uncovered new cell-type information for *risk tolerance, MDD, schizophrenia*, and *bipolar disorder*. These enrichments highlight unique cell-specific processes underlying these disorders which overlap in phenotypically and genetically similar phenotypes *schizophrenia* and *bipolar disorder*: the cell-types discovered in the conditioned *schizophrenia* GWAS overlap with those in the conditioned *bipolar disorder* GWAS. These findings recapitulate common therapeutic targets for these disorders. For example, inhibitory and GABAergic neuron transcriptomic profile enrichments were detected in the conditioned *MDD* GWAS and these are common targets of emerging therapeutic options (e.g., scopolamine, an antidepressant which blocks the M1 receptor of GABAergic interneurons in the medial prefrontal cortex (26); ketamine blocking the activation of somatostatin interneurons in PFC (27)) for *MDD* and *depressive symptoms* (26). Detection of these overlapping cell-type transcriptomic profile enrichments supports drug repurposing efforts in psychiatric disorders and related mental health conditions.

Using genome-wide information, we uncovered putatively causal relationships between many mental health outcomes. These analyses revealed well-known relationships between traits (e.g., *bipolar disorder, schizophrenia*, and *MDD*) but also elucidated several novel relationships involving brain imaging phenotypes. In particular we identified the *volume of the left subcallosal cortex* as a possible mediator of the relationships between several mental health outcomes (e.g., *extraversion, subjective well-being*, and *alcohol dependence*) which in turn demonstrate potential causal relationships with mood disorders which are commonly comorbid with alcohol dependence (28). This structural convergence may elucidate common disease mechanisms; however, these commonalities may be confounded by fine-grained nuances of the relationship between brain structure and mental health and disease. The LCV method used to identify these causal relationships does not support multivariable analyses nor does it employ sensitivity tests to detect horizontal pleiotropy (i.e., a SNP is associated with both phenotypes through separate mechanisms) or effect-size outlier SNPs. These observations likely confound our causal inferences and require more robust testing with traditional Mendelian randomization methods suited to accommodate weak genetic instruments (i.e., those SNPs not strongly associated with either phenotypes of interest, typically with association p-values greater than 5×10^−8^) (29-31).

Certain relationships regarding mental health outcome conditioning that might have been expected, were not observed in our study. Intellectual abilities are genetically correlated with *ASD* and *ADHD* and disabilities therein often co-occur with *ASD* and/or *ADHD* diagnoses (32, 33). According to the Diagnostic and Statistical Manual of Mental Disorders (5^th^ Edition, DSM-5), diagnosis of intellectual disability or global developmental delay must be eliminated as possible explanations of ASD symptoms prior to making a formal ASD diagnosis. We had hypothesized that after conditioning with the effects of EDU phenotypes, these psychiatric disorders might demonstrate notable changes in their underlying biology, but this was not the case. This lack may suggest that ADHD and ASD diagnosis criteria robustly capture elements unique to the disorders rather than those shared with EDU/SES phenotypes. In other words, ascertainment of cases at the extreme ends of spectrum disorders (34, 35) reliably capture trait specific biology with minimal phenotype confounding from shared effects with EDU and SES.

### Education and Socioeconomic Status

EDU and SES phenotypes were generally robust to the effects of shared genetic etiology with mental health outcomes. When conditioned with individual mental health outcomes, we detected relatively few changes to the predicted underlying biology of EDU and SES phenotypes. Only when EDU and SES phenotypes were conditioned with several mental health outcomes did we observed changes in h^2^ and underlying biology. The phenotypes *educational attainment, income*, and *self-rated math ability* maintained significant h^2^ after conditioning with all mental health outcomes. Conversely, the SNP-based observed-scale h^2^ of *cognitive performance, highest math class*, and *deprivation index* disappeared after conditioning with all mental health outcomes. When assessing the relationship between EDU and SES phenotypes, we revealed *educational attainment* as a driving factor linking EDU and SES phenotypes. Furthermore, we uncovered an independent contribution of *income* to a common factor with *educational attainment* and *self-rated math ability*. Based on recent work of Morris, et al. to uncover why EDU and SES phenotypes are related to one another, these observations point to *educational attainment* as a mediator of the genetic and phenotypic correlations between EDU and SES.

Tissue and cell-type transcriptomic profile analyses of EDU, SES, and mental health outcome phenotypes highlighted differences in cortical and cerebellar tissue enrichment. Though not significantly decreased in all phenotypes after conditioning, the bidirectional changes in cerebellar and cortical tissue enrichment (i.e., EDU/SES conditioned with mental health outcomes and mental health outcomes conditioned with EDU/SES) highlight the importance of these brain regions and their shared transcriptional regulation in human mental health and disease (36). Furthermore, this observation of cerebellar and cortical tissue changes support the common psychopathology factor (a p-factor) studied extensively in recent mental health and disease research (37). Genetic risk and structural brain imaging changes have been identified underlying this p-factor (37).

### Limitations

Our study has three primary limitations. First, we did not select independent genetic correlates from the mental health outcome phenotypes with which to condition the EDU and SES phenotypes. Due to high genetic correlation between mental health outcomes, this approach may have introduced bias in our reporting of which EDU and SES phenotypes were robust to shared genetic etiology with all mental health outcomes. This potential over-conditioning likely drove our results towards the null (e.g., non-significant h^2^) and therefore, we have not reported gene set, tissue transcriptomic profile enrichment, cell-type transcriptomic profile enrichment, or GenomicSEM loadings for EDU/SES traits where there might have been over-conditioning. For this reason, our results do not imply that, for example, *educational attainment* is a more powerful or specific EDU phenotype than *cognitive ability*. Second, it has recently been demonstrated that the origin of phenotypic and genetic correlations between EDU and SES phenotypes may be driven by dynastic effects and/or assortative mating acting independently or in concert (6). Dynastic effects describe a condition where offspring inherit phenotype-associated genetic risks and phenotype-associated environmental risks. Assortative mating exists when mate pairs are non-randomly chosen based on certain attributes. We hypothesize that the dynastic and assortative mating events described between EDU and SES phenotypes (6) may also appear in phenotypic and genetically correlated EDU, SES, and mental health outcome pairs. Future studies will be required to describe how these evolutionary and social pressures influence the correlative and causal relationships uncovered here (e.g., OCD→anorexia nervosa after conditioning with the effects of *educational attainment, income*, and *deprivation index*). Third, The UK Biobank is considered a generally healthy cohort not enriched for any trait or disorder of interest. To our knowledge, the brain imaging GWAS (performed on a subset of UK Biobank participants) used here were adjusted for variables related to blood pressure, height, weight, and bone mineral composition but are not adjusted for substance-related (recreational or prescribed) or psychiatric variables. The presence of these variables in sufficient quantities among those brain imaging participants could alter brain volumes affecting the results of the genetic analyses conducted.

## Conclusions

By conditioning mental health outcomes for the shared genetic etiology with EDU and SES phenotypes, this study elucidates novel biology and causal relationships between phenotypes. These biological mechanisms, cell-types, tissue-types, and causal trait pairs could not have been detected without adjusting the effects of EDU and SES. This study highlights how the pervasive effects of EDU and SES may mask underlying biology of mental health outcomes in support of multitrait analyses of GWAS to enable trait-specific discoveries.

## Materials and Methods

An overview of all materials, methods, and key findings from this investigation of the genetic overlap between EDU, SES, and mental health outcome phenotypes is shown in a flow diagram in Fig. S1.

### Trait Description and Selection

Four EDU (*educational attainment, highest math class, self-rated math ability*, and *cognitive performance*) and two SES phenotypes (*household income* and *Townsend deprivation index*) from the Social Science Genetic Association Consortium (SSGAC), UK Biobank (UKB), and 23&Me were used in this study to condition mental health outcomes. These unconditioned phenotypes are characterized on the level of heritability, tissue transcriptomic profile enrichment, and cell-type transcriptomic profile enrichment in Fig. S1.

Mental health outcomes from the Psychiatric Genomics Consortium (PGC), SSGAC, Genetics of Personality Consortium (GPC), UKB, and UKB Brain Imaging Genetics (UKB BIG) were selected based on their genetic correlation with EDU and SES phenotypes (Table S1 and Figs. 1 & S2). To focus our analyses, we predetermined that mental health outcomes would be included if (1) they had significant heritability (h^2^), and (2) they were genetically correlated with 2/4 EDU and 2/2 SES phenotypes. It is recommended that each phenotype in a genetic correlation pair have h^2^ z-scores ≥ 4 (18) but mtCOJO (see below) only requires significant heritability estimates. For this reason, we have relaxed the h^2^ suggestions for genetic correlation analyses with respect to trait inclusion. Genetic correlation estimates should be interpreted in light of this relaxed h^2^ criteria.

### Conditioning

Conditioning was performed in Genome-wide Complex Trait Analysis (GCTA) using the mtCOJO feature using the 1000 Genomes Project European ancestry linkage disequilibrium reference panel (17). For case-control GWAS summary statistics, odds ratios and corresponding standard error were converted to log-odds and corresponding standard error.

Causal estimates within mtCOJO were calculated using Generalized Summary-data-based Mendelian Randomization (GSMR). In our analyses of mental health outcomes conditioned with the effects of EDU and SES phenotypes, each MR causal inference was performed using genome-wide significant SNPs in the exposure (EDU/SES trait). To test how mental health outcomes influence EDU and SES phenotypes, we relaxed this SNP inclusion threshold where necessary (e.g., when UKB BIG phenotypes served as the exposure phenotype) such that at least two SNPs were included in the causal inference.

### Heritability and Genetic Correlation

Observed-scale h^2^ was calculated for each original and conditioned GWAS using the Linkage Disequilibrium Score Regression (LDSC) method with 1000 Genomes Project European reference population (18).

### Gene-set, Tissue Transcriptomic, and Cell-type Transcriptomic Profile Enrichment

Original and conditioned GWAS were used as standard input for Multi-marker Analysis of GenoMic Annotation (MAGMA v1.06) implemented in FUnctional Mapping and Annotation (FUMA) v1.3.3c with the following parameters: genome-wide significance p < 5×10^−8^, minor allele frequency ≥ 0.01, and LD blocks merged at < 250kb for LD r^2^≥0.6 with lead variant (22, 38).

SNPs underlying each phenotype of interest were mapped to genes within 10kb physical proximity using FUMA (38). Mapped genes were assessed using the gene-set enrichment feature of FUMA, and gene ontology enrichment analysis with ShinyGO (39).

Tissue transcriptomic profile enrichment was performed relative to the GTEx v7 53 tissue types with the default 0kb gene window.

Cell-type transcriptomic profile enrichments were performed using eleven human specific transcriptomic profile datasets related to the brain (22): PsychENCODE_Developmental, PsychENCODE_Adult, Allen_Human_LGN_level 1, Allen_Human_MTG_level1, DroNc_Human_Hippocampus, GSE104276_Human_Prefrontal_cortex_all_ages, GSE104276_Human_prefrontal_cortex_per_ages, GSE67835_Human_Cortex, GSE67835_Human_Cortex_woFetal, Linnarson_GSE101601_Human_Temporal_cortex, and Linnarson_GSE76381_Human_Midbrain. Cell-type transcriptomic profiles were assessed in three ways as per the FUMA analysis pipeline. (1) enrichment of cell-type transcriptomic profiles within each selected data set, (2) within data set conditionally independent cell-type transcriptomic profile enrichments, and (3) across data set cell-type transcriptomic profile enrichments.

For analyses within data sets, step-wise conditional significance is evaluated for each cell type in a data set against the p-values for all other cell-types in that same data set. The output from these analyses identify cell types within a data set whose transcriptomic profiles are enriched in a given GWAS independently of the signal from all other cell type transcriptomic profiles in the same data set.

Using within-data-set conditionally independent cell-types identified above, cross-data-set analysis identifies cell-type transcriptomic profiles enriched in a given GWAS independent of all other cell-types from all chosen data sets. Proportionally significant (PS) and conditionally independent cell-type pairs indicate that enrichment of these cell-types in a given GWAS are driven by independent genetic signals.

For a given pair of cell types, PS of cell type a given cell type b (PS_a,b_) ≥ 0.8 and PS_b,a_ ≥ 0.8 indicates independent genetic signals for cell types a. Interpretation of additional PS thresholds for each cell type in a given pair can be seen in detail (https://fuma.ctglab.nl/tutorial#celltype) or in Watanabe, et al. (22).

### Latent Causal Variables

LCV is a method for inferring genetic causal relationships between trait pairs using GWAS summary data using effect size estimates or z-scores (24). LCV modeling was implemented in R using the 1000 Genomes Project Phase 3 European reference panel. As recommended, GWAS summary data were filtered to include only SNPs with minor allele frequencies greater than 5% and the major histocompatibility region was removed due to its complex linkage disequilibrium structure. Note that genetic correlation does not imply that shared genetic risks between traits are causal. The LCV model output distinguishes whether genetic correlations support genetic causation and the degree to which (i.e., the genetic causality proportion; gĉp) genetic risk for trait 1 is causal for trait 2. LCV gĉp estimates were only interpreted for trait pairs where both traits exhibit LCV-calculated h^2^ z-scores ≥ 7.

### Statistical Considerations

Z-tests were used to determine differences in heritability, SNP effects, gene-set enrichments, tissue transcriptomic profile enrichments, cell-type transcriptomic profile enrichments, GenomicSEM loadings, and LCV estimates between conditioned and unconditioned GWAS.

## Data Availability

All data used in this study are available for public access and download for scientific purposes.

https://www.med.unc.edu/pgc/results-and-downloads/

https://www.thessgac.org/

http://www.tweelingenregister.org/GPC/

http://big.stats.ox.ac.uk/

## General

We would like to thank the research participants and employees of 23andMe Inc for making this work possible.

## Author contributions

FW and RP conceived the analyses; FW performed analyses; FW, GP, and RP interpreted the data; TL, JK, and JG provided critical analytic and data interpretation feedback; FW drafted the manuscript; FW, GP, TL, JK, JG, and RP edited and approved final manuscript.

## Data and materials availability

All data and analysis materials used in the study are publicly available for download.

## Figure and Figure Legends

**Table 1. Causal inferences detected after multiple testing correction.** Significant causal relationships detected between mental health outcomes using latent causal variable analyses. For each significant causal estimate, all conditioned causal estimates between that phenotype pair are provided, highlighting at least nominally significant causal inferences after conditioning with education and socioeconomic status phenotypes that could not be detected in the original unconditioned trait pair.

**Submitted as a separate file**.

## Supplementary Materials

### Supplementary Results

#### Trait Inclusion

Left and right insular cortex brain imaging phenotypes have been treated as differentially genetically correlated with EDU and SES, respectively. (1) Though not included in the conditioning experiments involving EDU phenotypes, the right insular cortex was nominally significantly genetically correlated with educational attainment (r_g_ = −0.118, p = 0.044) and (2) though not included in the conditioning experiments involving SES phenotypes, the left insular cortex was nominally significantly genetically correlated with income (r_g_ = 0.183, p=0.004).

#### Gene-set Enrichment Differences

Using MAGMA (38), we tested for differences between the gene set enrichments of original and conditioned mental health outcome GWASs. Gene set enrichments for each mental health outcomes were robust to the effects of conditioning with EDU and SES phenotypes. No significant changes were observed in gene set enrichments underlying the GWAS of mental health outcomes.

**Fig. S1.**
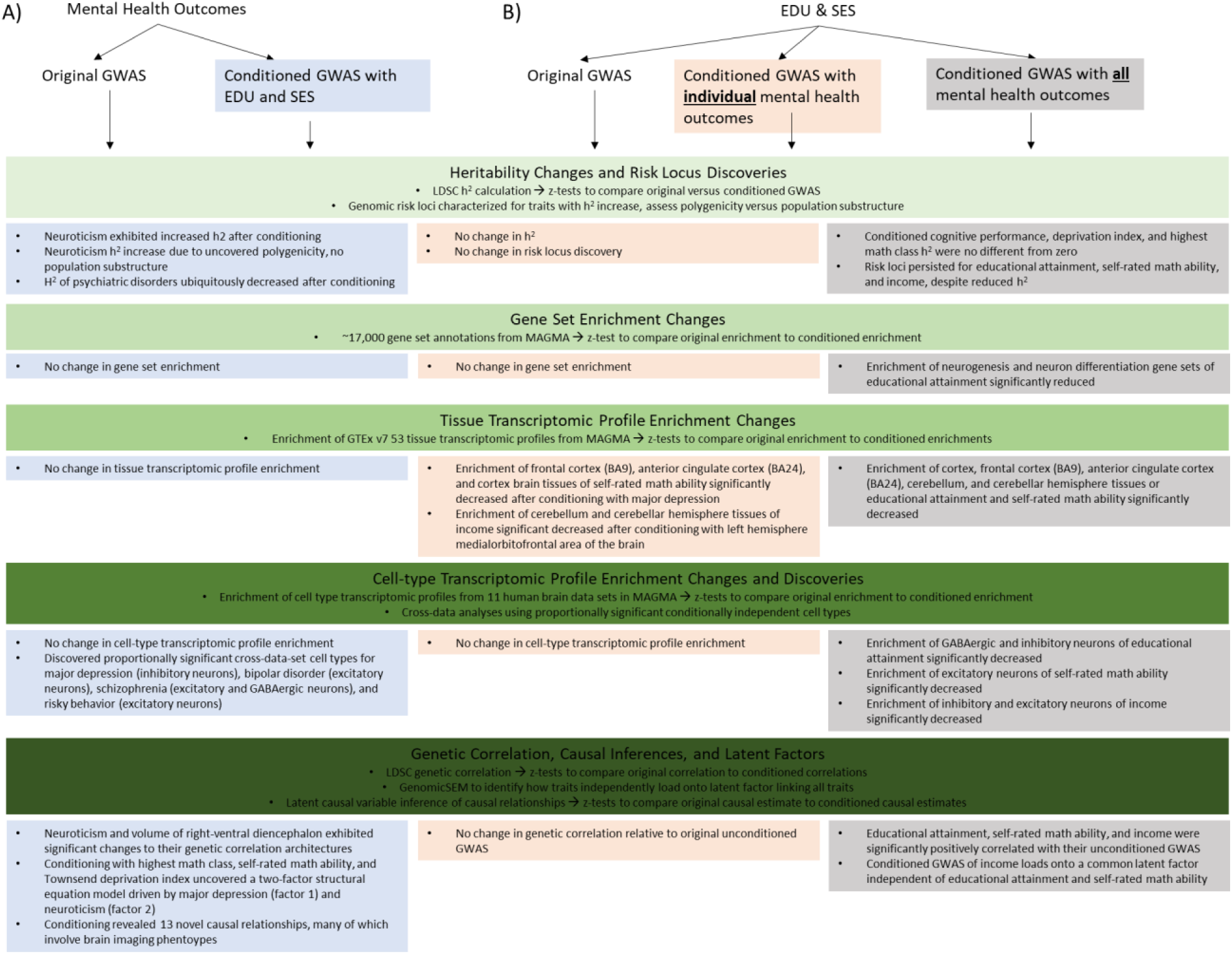
Analytic pipeline. The overall analysis plans and methods used (green boxes) and key results from each phase of analysis for **(A; blue boxes)** mental health outcomes conditioned with education and socioeconomic status phenotypes and **(B)** education and socioeconomic status phenotypes conditioned with individual mental health outcomes (orange boxes) and all mental health outcomes (grey boxes).

**Fig. S2.**
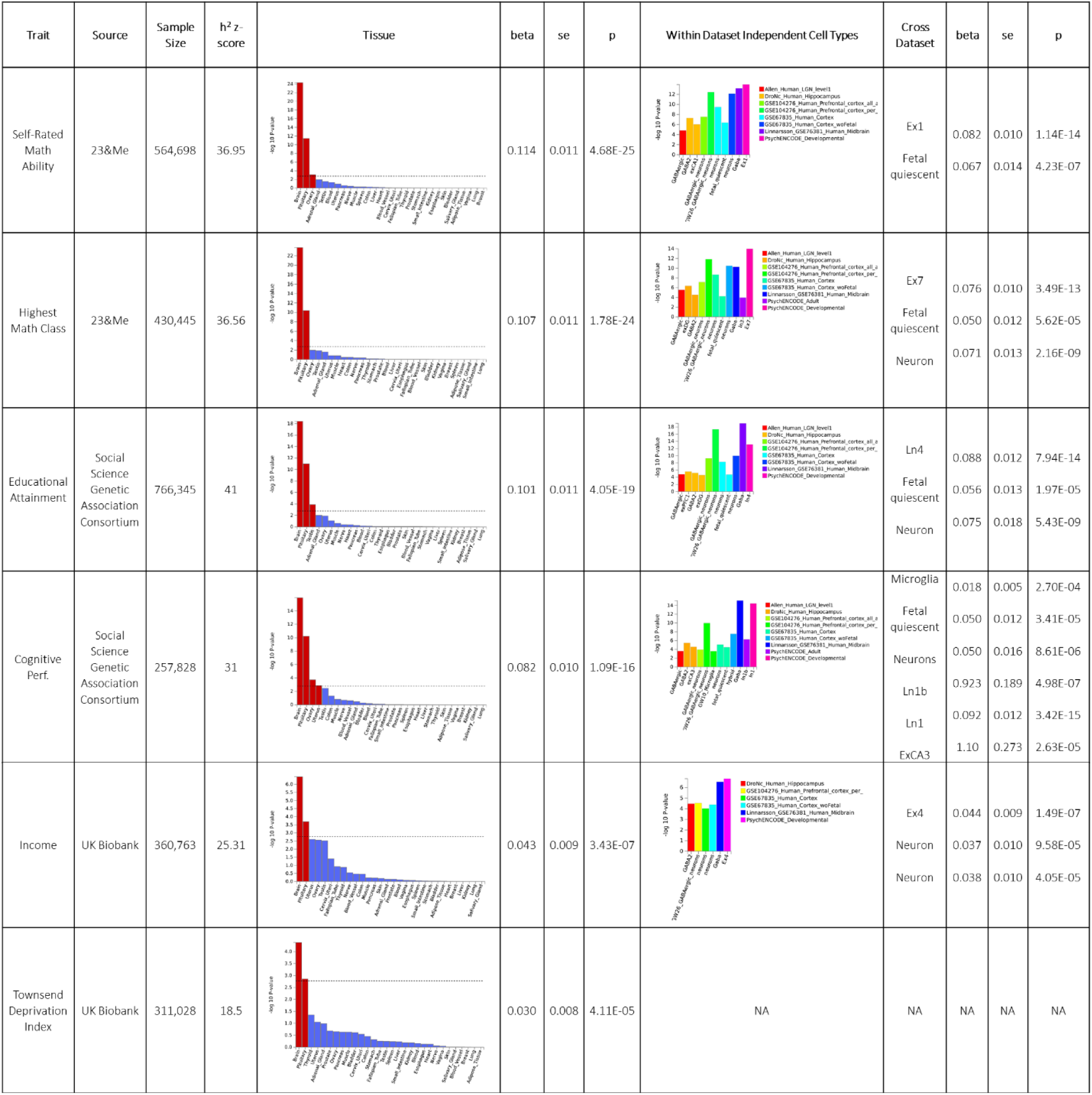
Description of education and socioeconomic status phenotypes. Summary level data for education and socioeconomic status phenotypes on the level of phenotype heritability, tissue transcriptomic profile enrichment, and cell-type transcriptomic profile enrichment.

**Fig. S3.**
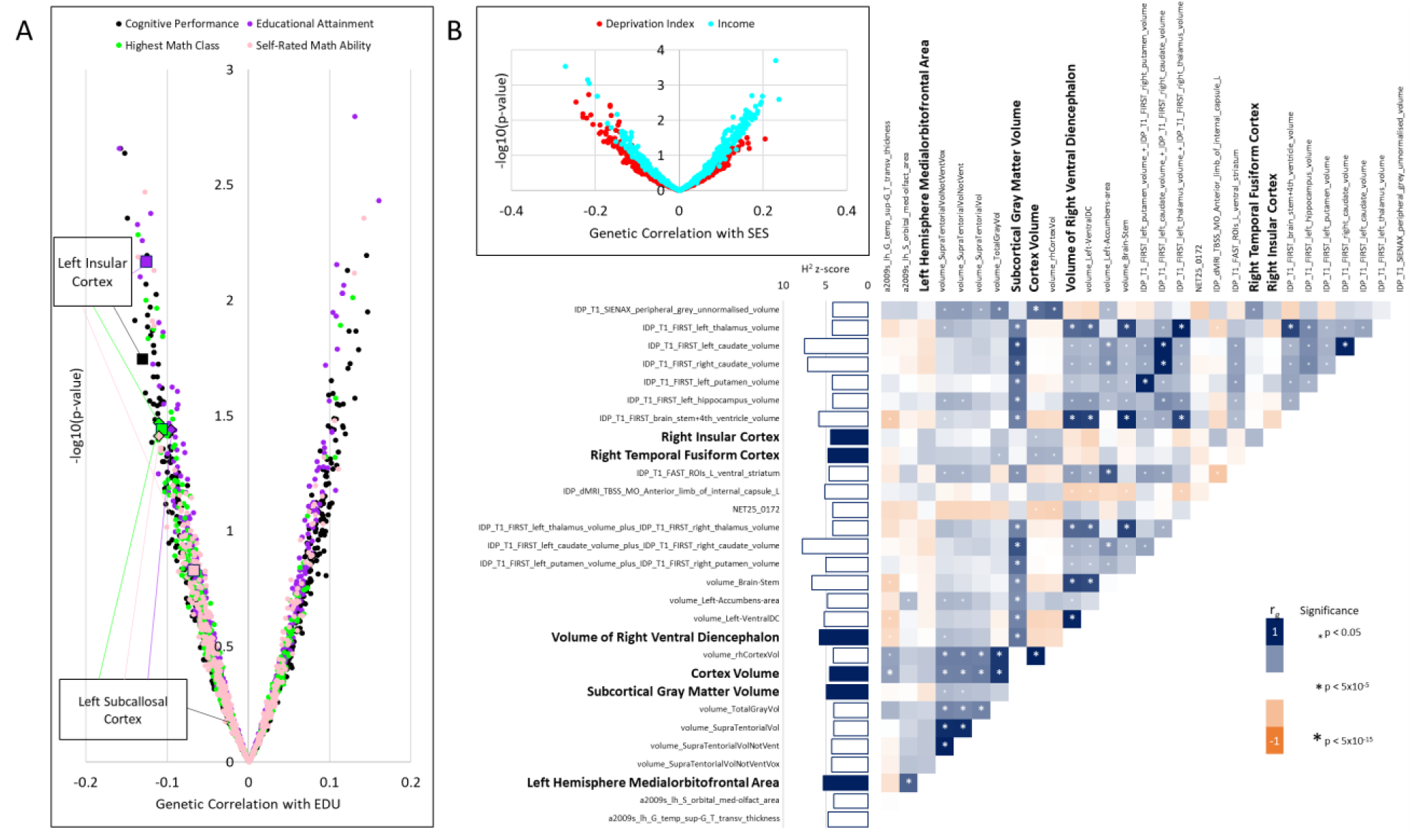
Selection of brain imaging phenotypes. **(A)** Brain imaging phenotypes genetically correlated with education phenotypes: educational attainment, highest math class, self-rated math ability, and cognitive performance. **(B)** Brain imaging phenotypes nominally genetically correlated with socioeconomic status phenotypes: household income and Townsend deprivation index. Heatmap shows genetic correlations between socioeconomic status genetic correlates and their respective per-trait heritability z-scores. Six bolded brain imaging phenotypes were selected for conditioning experiments due to their heritability estimate relative to strongly genetically correlated traits.

**Fig S4.**
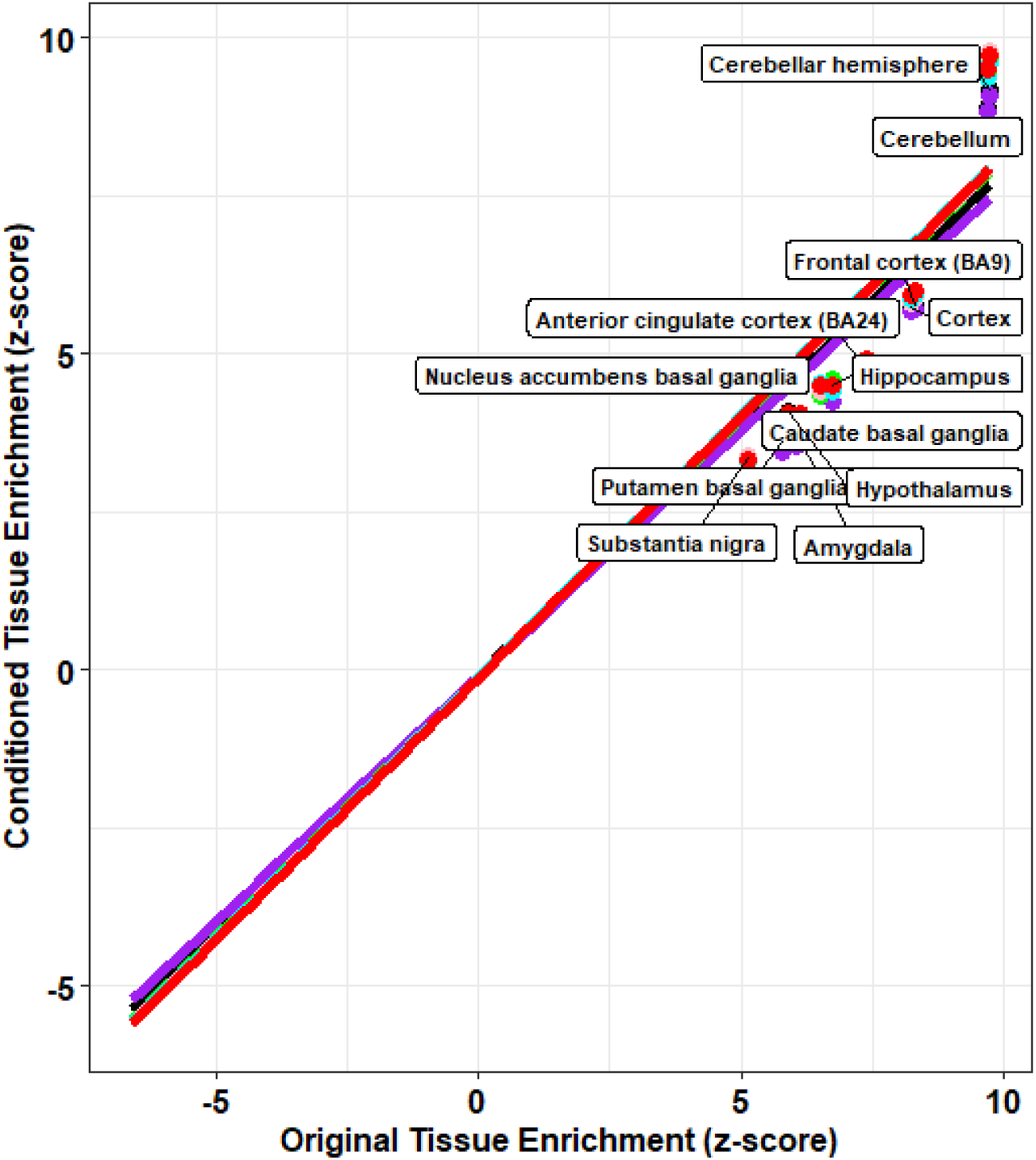
Tissue changes in schizophrenia GWAS. Robust linear regression between original and conditioned tissue transcriptomic profile enrichments in the schizophrenia GWAS. The schizophrenia GWAS was conditioned with education and socioeconomic status phenotypes: *educational attainment* (purple regression line), *highest math class* (green regression line), *self-rated math ability* (pink regression line), *cognitive performance* (black regression line), *income* (cyan regression line), and *Townsend deprivation index* (red regression line). Tissues are labeled if conditioning with education and socioeconomic status significantly reduced their enrichment in the schizophrenia GWAS.

**Fig. S5.**
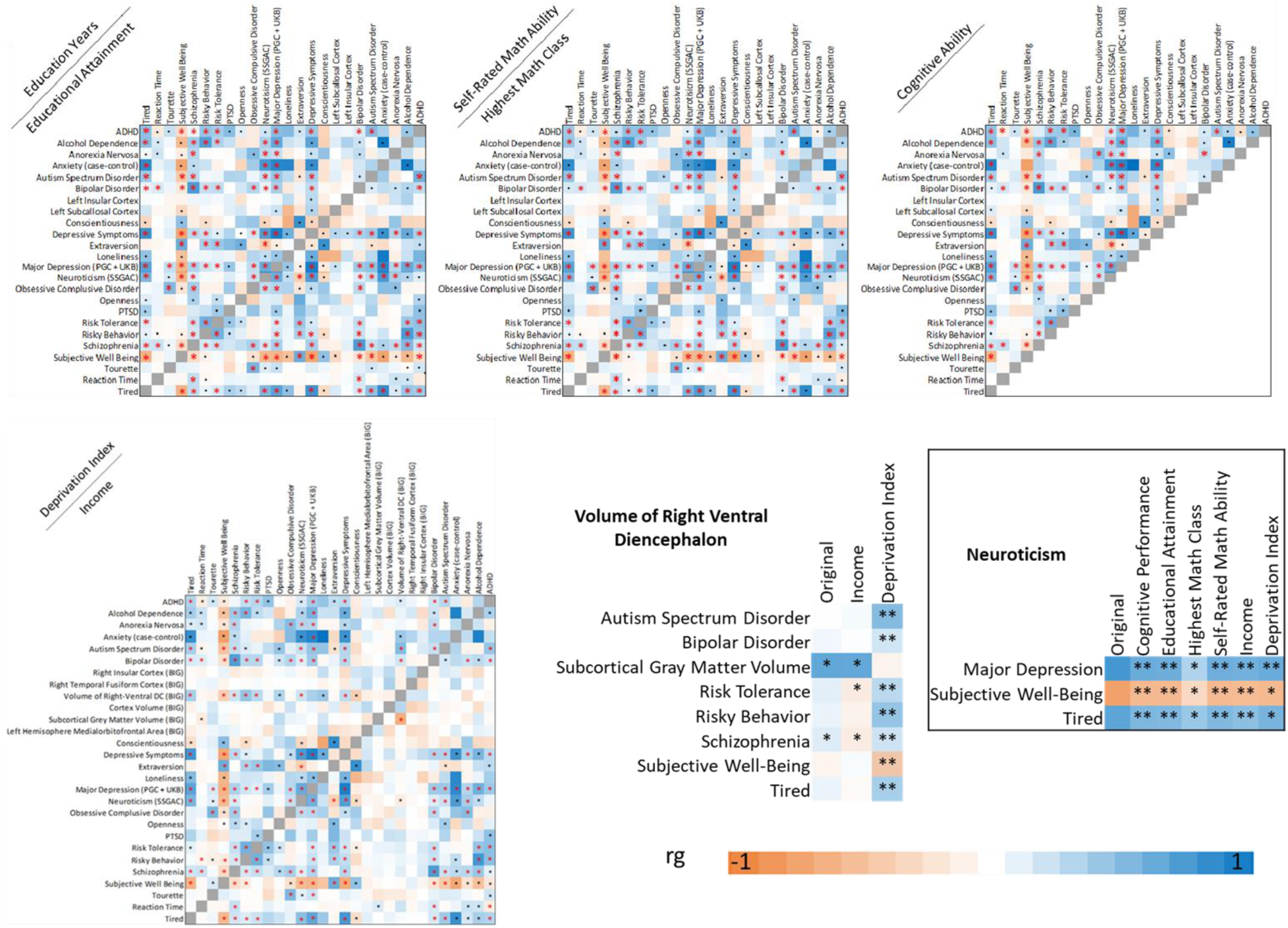
Mental health outcome genetic correlations. Genetic correlation architecture of all mental health outcomes shown as heatmaps. Red asterisks indicate genetic correlations surviving Bonferroni correction, single black asterisks indicate at least nominally significant genetic correlations, and double black asterisks indicate genetic correlations that are significantly different than their unconditioned equivalent.

**Fig. S6.**
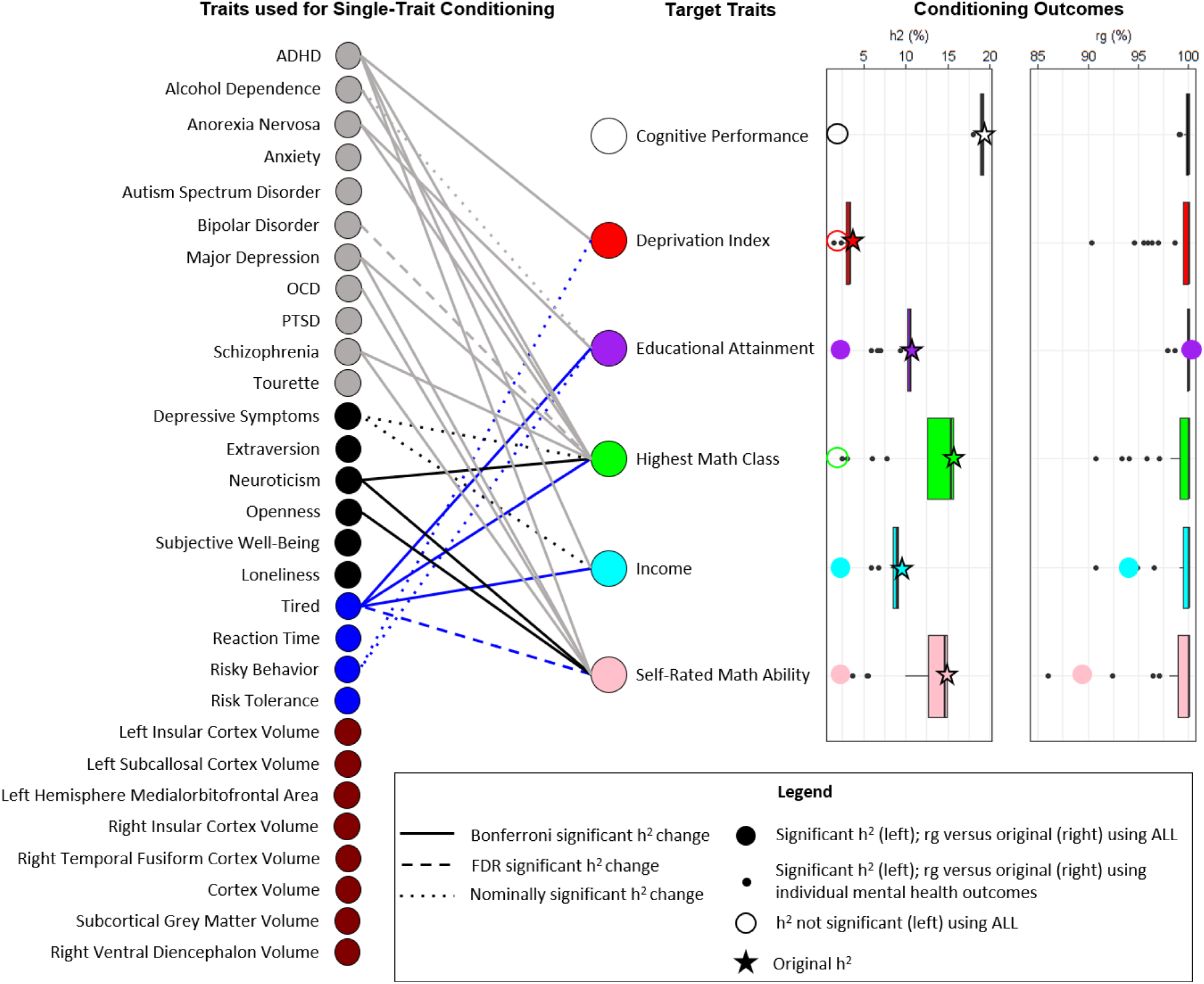
Heritability and genetic correlation of education and socioeconomic status phenotypes.

**Fig S7.**
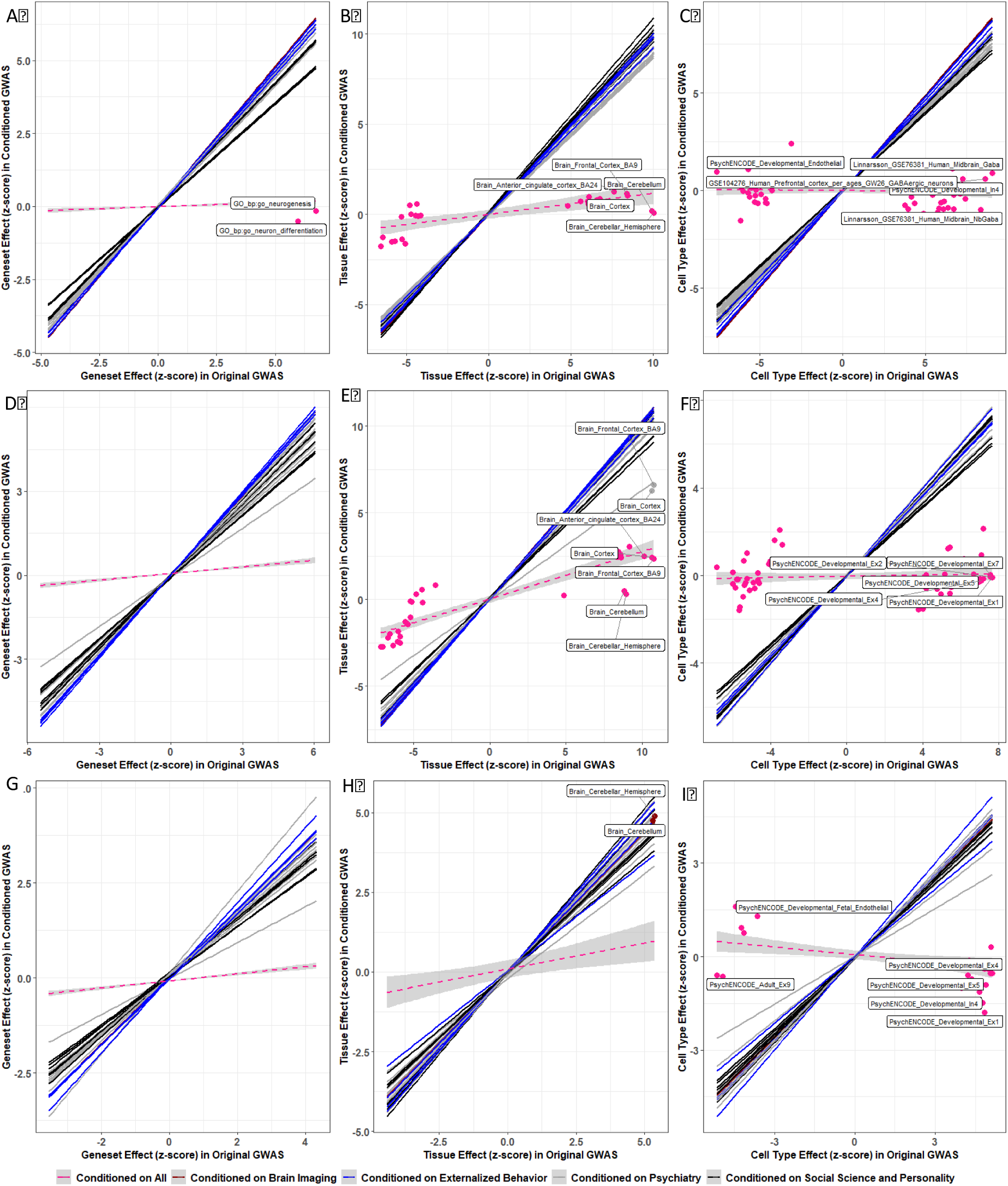
Gene set, tissue transcriptomic profile, and cell-type transcriptomic profile enrichment changes. Each panel shows the robust linear regression between original gene set **(A, D, G)**, tissue transcriptomic profiles **(B, E, H)**, and cell-type transcriptomic profiles **(C, F, I)** and conditioned enrichments for *educational attainment* **(A-C)**, *self-rated math ability* **(D-F)**, and *income* **(G-I)**. For visual clarity, standard errors around each linear regression are shown for traits resulting in a significant change of enrichment and for each trait conditioned on all mental health outcomes (pink dashed regression lines). Data points indicate annotations with significant different enrichments after conditioning; the top five differentially enriched annotations are labeled.

**Fig. S8.**
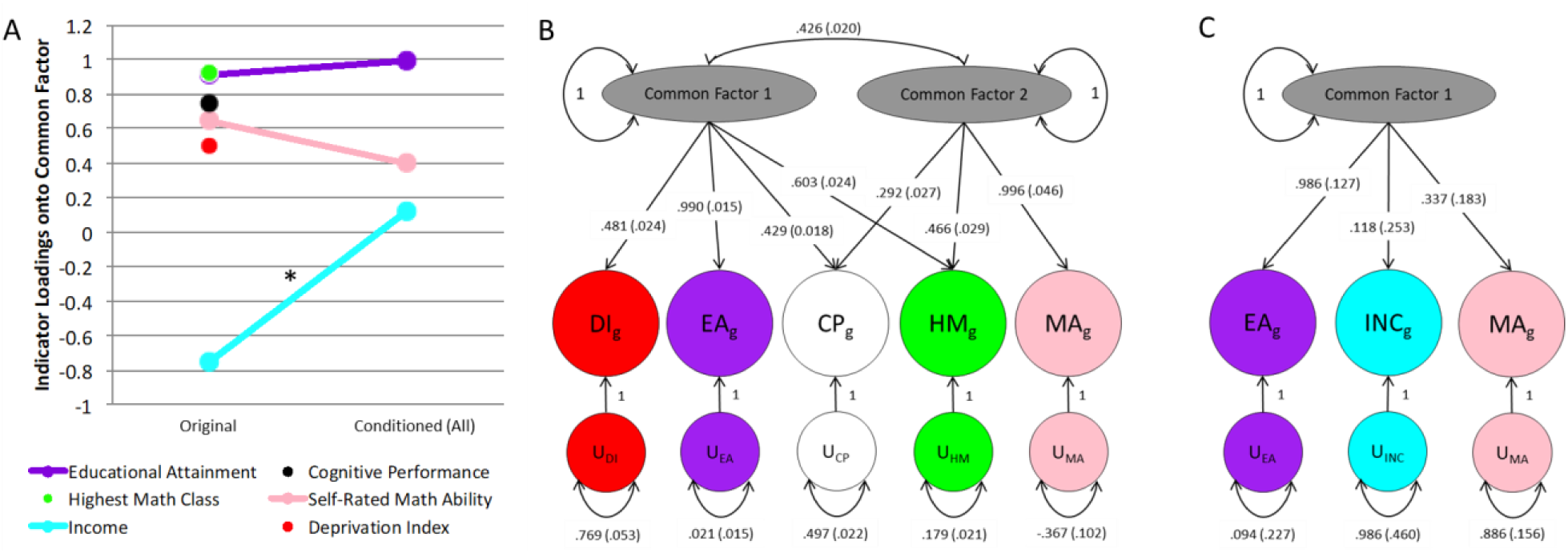
Summary of Genomic Structural Equation Modeling (GenomicSEM). **(A)** SEM of education (EDU) and socioeconomic status (SES) phenotypes assuming all phenotypes load onto a single common factor. The same model with significantly heritably conditioned phenotypes is shown. Asterisks indicate significant changes in loading value. **(B)** Confirmatory factor analysis of unconditioned EDU and SES proxies. **(C)** Confirmatory factor analysis of EDU and SES proxies conditioned with all mental health outcomes.

**Table S1.** Description of mental health outcomes assessed.

| Trait | Source | Sample Size | H<sup>2</sup> % (z-score) |
| --- | --- | --- | --- |
| Autism Spectrum Disorder | PGC | 18,382 cases and 27,969 controls | 19.41 (11.55) |
| Attention Deficit Hyperactivity Disorder | PGC | 20,183 cases and 35,191 controls | 22.81 (15.41) |
| Schizophrenia | PGC | 36,989 cases and 113,075 controls | 23.22 (25.24) |
| Post-traumatic Stress Disorder | PGC | 2,424 cases and 7,113 controls | 4.86 (2.26) |
| Anorexia Nervosa | PGC | 3,495 cases and 10,982 controls | 24.03 (6.29) |
| Alcohol Dependence | PGC | 8,485 cases and 20,272 controls | 5.08 (4.38) |
| Major Depressive Disorder | PGC + UKB | 246,363 cases and 561,190 controls | 3.75 (25) |
| Bipolar Disorder | PGC | 20,352 cases and 31,358 controls | 4.32 (12) |
| Tourette Syndrome | PGC | 4,819 cases and 9,488 controls | 35.62 (8.32) |
| Obsessive Compulsive Disorder | PGC | 2,688 cases and 7,037 controls | 34.56 (7.2) |
| Anxiety (Case-Control) | ANGST | 5,761 cases and 11,765 controls | 7.26 (2.49) |
| Subjective Well-Being | SSGAC | 298,420 | 2.5 (12.5) |
| Neuroticism | SSGAC | 170,911 | 9.41 (14.26) |
| Depressive Symptoms | SSGAC | 161,460 | 4.72 (12.76) |
| Risk Tolerance | SSGAC | 466,571 | 5.13 (23.32) |
| Risky Behavior | SSGAC | 315,894 | 11.09 (26.4) |
| Openness | GPC | 17,375 | 9.27 (4.14) |
| Conscientiousness | GPC | 17,375 | 6.32 (2.47) |
| Extraversion | GPC | 63,991 | 5.19 (6.33) |
| Loneliness | Gao, et al. 2017 | 2,853 cases and 4,583 controls | 0.3 (2.14) |
| Reaction Time | CCACE/UKB | 330,069 | 8.13 (27.1) |
| Tiredness | CCACE/UKB | 108,976 | 5.84 (21.63) |
| Left Insular Cortex | BIG | 8,428 | 23.13 (4.18) |
| Right Insular Cortex | BIG | 8,428 | 21.76 (4.20) |
| Left Subcallosal Cortex | BIG | 8,428 | 22.96 (4.34) |
| Right Temporal Fusiform Cortex | BIG | 8,428 | 26.03 (4.51) |
| Volume of Right-Ventral Diencephalon | BIG | 8,428 | 32.83 (5.41) |
| Cortex Volume | BIG | 8,428 | 22.36 (4.47) |
| Subcortical Grey Matter Volume | BIG | 8,428 | 27.58 (4.57) |
| Left Hemisphere Medialorbitofrontal Area | BIG | 8,428 | 23.11 (4.83) |

